# Sustainability of Lean interventions in public hospitals after a national quality improvement programme: a multicentre mixed-methods study

**DOI:** 10.64898/2026.08.08.26359963

**Authors:** Breno Douglas Dantas Oliveira, Marco Saavedra Bravo, Wendell Guilherme Ribeiro do Prado, Paula de Almeida Ruiz, Carina Tischler Pires

**Author notes:** **Corresponding author:** Breno Douglas Dantas Oliveira, Hospital Sírio-Libanês, São Paulo, Brazil.

## Abstract

**Objectives:** To evaluate the sustainability of Lean Healthcare practices after the implementation phase of a national quality improvement programme and to identify organisational factors associated with maintaining results over time.

**Design:** Multicentre cross-sectional study with a mixed-methods approach.

**Setting:** Twelve public and philanthropic hospitals in Brazil participating in Phase 2 of the Lean in Emergency Departments Project.

**Participants:** Key respondents in managerial or leadership roles from participating hospitals (response rate: 75.0%).

**Outcome measures:** Sustainability of Lean practices and organisational readiness, assessed through a structured survey and triangulated with operational indicators collected across successive implementation cycles at hospital level.

**Results:** During one year of structured follow-up, 66.7% of respondents reported maintenance of Lean practices; this decreased to 33.3% after the end of structured follow-up. Although 66.7% considered professionals capable of maintaining results, only 58.3% positively evaluated institutional structure, indicating a discrepancy between individual capacity and organisational readiness. Operational indicators showed heterogeneous behaviour across hospitals, with no consistent pattern of sustained improvement. Qualitative analysis identified professional and managerial turnover, formal governance structures, and continuous monitoring as key factors associated with sustainability.

**Conclusions:** The sustainability of Lean Healthcare practices is more strongly associated with institutional capacity to embed and sustain changes over time than with isolated individual training. Quality improvement programmes should incorporate structured strategies for the post-implementation phase.

**STRENGTHS AND LIMITATIONS OF THIS STUDY:**

- This multicentre study combines survey data with longitudinal operational indicators, providing a comprehensive assessment of the sustainability of Lean Healthcare practices.
- The mixed-methods design enabled triangulation between quantitative and qualitative findings, strengthening the robustness of the results.
- The study reflects real-world implementation across public and philanthropic hospitals within a national quality improvement programme.
- The use of self-reported data from a single key respondent per hospital may introduce perception bias and social desirability bias, and may not fully capture the diversity of institutional perspectives on sustainability outcomes.
- The cross-sectional design limits causal inference regarding the mechanisms underlying sustainability.

## 1. INTRODUCTION

Emergency department overcrowding and prolonged hospital length of stay represent persistent challenges in contemporary healthcare systems, directly impacting care quality, patient safety, and hospital operational efficiency. This phenomenon is not restricted to the emergency setting but reflects structural and organisational limitations throughout the entire hospital care pathway, especially in admission processes, bed management, and hospital discharge.^1,2^

Various studies have demonstrated that delays in bed availability, often translated by the phenomenon of boarding, are associated with worse clinical outcomes, increased length of stay, and greater resource utilisation.^2,3^ In this context, interventions aimed at improving patient flow and optimising hospital capacity have been prioritised in different healthcare systems, with emphasis on Lean Healthcare approaches, which propose waste reduction, process standardisation, and strengthening of data-driven management.^4–8^

Although evidence indicates that Lean interventions can produce significant improvements in hospital operational indicators, important gaps remain related to the variability of observed effects and, mainly, to the sustainability of results over time. Many studies focus on the initial implementation phase, with little emphasis on the continuity of gains after structured support ends, especially in real-world contexts of complex, large-scale healthcare systems.^9,10^

In this scenario, it is crucial to investigate not only the initial effectiveness of interventions, but, above all, the factors associated with sustaining results over time and process stability, considering the influence of the institutional environment and governance strategies adopted.^11^

This study aimed to evaluate whether Lean interventions implemented through a national programme are sustained over time, and to identify organisational factors associated with their sustainability. Operational indicators related to hospital flow and bed management were analysed to support the interpretation of sustainability outcomes.

## 2. METHODS

### 2.1 Study Design

This is a multicentre cross-sectional survey with a mixed-methods approach, complemented by longitudinal operational data, conducted to evaluate the sustainability of Lean Healthcare practices and tools following implementation of a structured national quality improvement programme. The cross-sectional design was based on a single time-point survey, with longitudinal operational data used for triangulation purposes. The study was conducted and reported in accordance with the Strengthening the Reporting of Observational Studies in Epidemiology (STROBE) guidelines.

### 2.2 Context

The study was conducted within the Unified Health System (SUS), within the scope of the Lean in Emergency Departments Project, a national initiative conducted by the Ministry of Health in partnership with Hospital Sírio-Libanês, through PROADI-SUS. The project aims to reduce overcrowding in Brazilian public and philanthropic hospitals and improve patient flow through the application of Lean Healthcare principles.

The intervention is structured in sequential phases. Phase 1 corresponds to an intensive implementation period lasting approximately 12 months, involving on-site and remote support, aimed at improving emergency department care flow, reducing length of stay, and optimising hospital bed utilisation.

Phase 2, the focus of this study, is directed at hospitals previously exposed to Phase 1 and also has a duration of approximately 12 months. It is characterised as a period of consolidation of Lean practices and strengthening of organisational processes, with emphasis on bed management, admission and discharge flows, and operating theatre efficiency. During this phase, approximately 12 on-site visits are conducted interspersed with remote meetings (inter-visits), with an average duration of approximately 16 hours per on-site visit and approximately 1.5 hours per remote meeting. Additionally, there was a subsequent period of remote monitoring, with monthly virtual meetings over six months. This arrangement allows evaluation of the incorporation of practices into the routine functioning of institutions and the maintenance of results over time.

Before implementation, hospitals faced challenges related to overcrowding, fragmentation of care processes, and inefficiency in patient flow, especially at the interface between emergency department, inpatient units, and operating theatre.

### 2.3 Intervention

The intervention evaluated in this study corresponds to the structured consolidation of Lean Healthcare practices during Phase 2 of the Lean in Emergency Departments Project.

Implementation was conducted through iterative improvement cycles based on the DMAIC model (Define, Measure, Analyse, Improve and Control), which guided the diagnosis of hospital demand and capacity, analysis of care processes, implementation of action plans, and continuous monitoring of results. In this context, the intervention involved: structured diagnosis of demand and capacity; development and implementation of action plans for improving care flows; application of Lean best practices in the operating theatre and inpatient units; and management and monitoring of operational indicators.

Actions were conducted by a pair comprising a consultant physician and Lean methodology specialist, combining on-site activities and remote follow-up throughout the implementation period. These lines of action can be understood, from an analytical perspective, in dimensions related to diagnosis, process standardisation, active management, and organisational governance.

A detailed representation of the implementation model is available in Supplementary Material 1. The intervention was conducted based on principles of continuous improvement and iterative cycles inspired by the action research approach, with continuous adaptation to the local specificities of each institution.

Key Lean Healthcare terms used throughout this manuscript are defined as follows for readers unfamiliar with the methodology. Huddle: a brief daily team meeting focused on operational priorities and problem identification. Kamishibai: a visual audit tool using cards to verify process adherence at the point of care. Gemba walk: a structured observation of work processes conducted at the site where care is delivered. Makigami: a detailed process mapping technique used to identify waste and inefficiencies in administrative and clinical workflows. 5S: a workplace organisation method based on five principles — sort, set in order, shine, standardise, and sustain.

### 2.4 Intervention Study

This study evaluated the sustainability of the intervention in the post-implementation period (Phase 2), considering the maintenance of practices, their incorporation into institutional functioning, and the stability of results over time. Sustainability was defined as a multidimensional construct, involving the continuity of practices, institutionalisation of processes, and organisational capacity to maintain improvements over time.

In this context, sustainability constituted the central analytical focus of the study, with operational indicators used as a triangulation strategy for findings, allowing comparison between respondent perception and the behaviour of care processes, without the intention of establishing causal relationships.

### 2.5 Participants

Eligible were Brazilian public and philanthropic hospitals that participated in Phase 2 of the Lean in Emergency Departments Project between 2018 and 2023. A total of 16 hospitals were invited to participate in the study, of which 12 responded to the questionnaire, corresponding to a response rate of 75.0%. The participant flow is illustrated in Figure 1. The four non-responding hospitals did not complete the questionnaire after three follow-up attempts and were excluded from the analysis.

**Figure 1.**
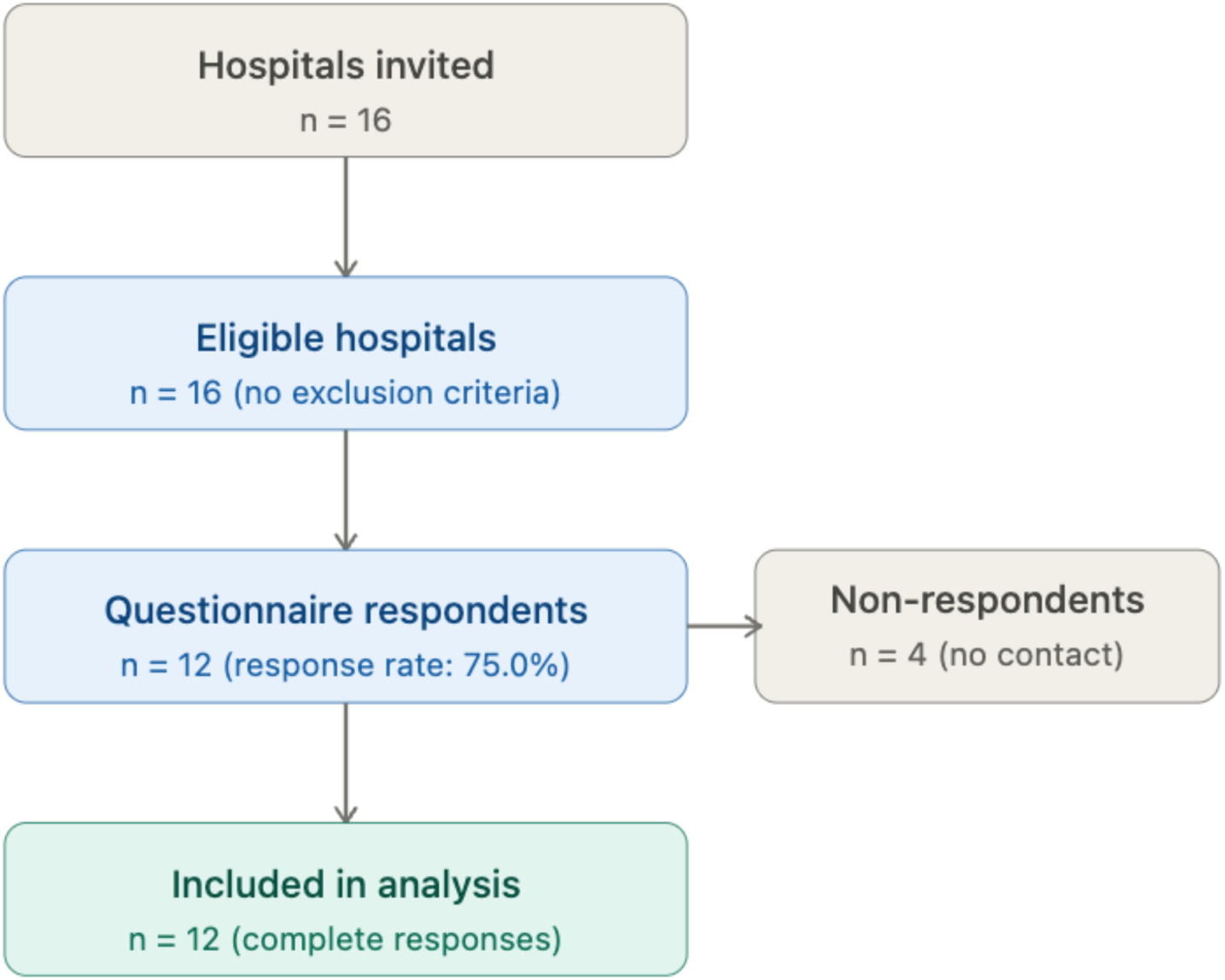
Participant flow diagram. Of 16 hospitals invited to participate, 12 completed the questionnaire (response rate: 75.0%) and were included in the analysis. Four hospitals did not respond after three follow-up attempts.

For each hospital, a single key respondent with a strategic or managerial role directly involved in project implementation was selected, with the aim of representing the institutional perspective on the continuity of Lean practices. Hospitals with incomplete responses or that could not be contacted were excluded from the analysis.

### 2.6 Data Collection

Data were collected through a structured, self-administered questionnaire, developed specifically for this study, based on the literature on Lean Healthcare and sustainability of health interventions. The instrument is presented in full in Supplementary Material 2, enabling study replication. The instrument was submitted to review by experts in hospital management and quality improvement, with focus on content validity and clarity of items.

The questionnaire was administered online through the Google Forms platform, with individual link sent by email to previously identified participants. Follow-up attempts were made at 7, 14, and 21 days for non-responding participants, with the aim of maximising the response rate. Data collection occurred between January and February 2026.

The instrument comprised 25 questions, including items on a five-point Likert scale, open-ended questions, and one dichotomous question. Responses were anonymous, without collection of information that would allow identification of participants.

### 2.7 Variables and Outcomes

The primary outcome of the study was the sustainability of Lean Healthcare practices. Variables were organised into four main domains: implementation and adoption of Lean practices; resistance and challenges during implementation; sustainability and maintenance of practices; and perceived impact and institutional readiness.

As complementary analytical elements, operational indicators from the project were considered, with the aim of supporting the interpretation of findings related to sustainability and allowing comparison between respondent perception and the behaviour of care processes. Operational indicators were defined according to standardised metrics, being considered: mean hospital length of stay (interval between admission and discharge); operating theatre efficiency (proportion of surgeries performed as scheduled); elective surgery cancellation rate (proportion of procedures cancelled on the scheduled day); and delay of first elective surgery (difference between scheduled time and actual start).

The operational definitions of these indicators followed definitions previously established in the Lean in Emergency Departments Project.

### 2.8 Data Analysis

Quantitative data were analysed using descriptive statistics, with presentation of absolute and relative frequencies, means and standard deviations, according to the nature of variables. Considering the number of participating units (n=12), inferential analyses were treated as exploratory. Quantitative analyses were performed using R software, version 4.x (R Foundation for Statistical Computing, Vienna, Austria).

Qualitative data from open-ended questions were analysed using reflexive thematic analysis, following the methodological framework proposed by Braun and Clarke.^12^ The textual corpus comprised 59 substantive text units extracted from seven open-ended questions from the questionnaire, corresponding to 1,691 words. All 12 respondents provided responses to the central open-ended questions (main challenges, strategies adopted, and suggestions for improvement). The analytical process was conducted in six stages: (1) floating reading of the corpus for familiarisation with the data; (2) generation of initial codes systematically from all material; (3) search for themes by grouping codes into potential categories; (4) review of themes in relation to the corpus and dataset; (5) final definition and naming of themes; and (6) production of the analytical report.

Coding was conducted independently by two researchers, with subsequent comparison and resolution of disagreements by consensus. Themes were derived inductively, guided by the content of responses, without imposition of prior theoretical structure. No computerised qualitative analysis software was used; analysis was conducted manually with support of spreadsheet for organisation of codes and themes.

Integration of quantitative and qualitative data was performed through convergent triangulation, comparing findings from the two components at the end of individual analyses, with the aim of identifying patterns of convergence, complementarity, or discrepancy. This process involved direct comparison between institutional perception reported in open-ended responses and Likert scale data and operational indicators, without a priori hierarchisation between components.

The researchers acknowledge their previous involvement in the Lean in Emergency Departments programme as a potential source of interpretive bias. To mitigate this, coding was conducted independently by two researchers, with explicit discussion of disagreements and close adherence to the data. Missing data were minimal and handled using complete-case analysis, as only fully completed questionnaires were included in the final analysis.

### 2.9 Patient and public involvement

Patients and the public were not directly involved in the design, conduct, reporting, or dissemination of this research. This study evaluated the sustainability of organisational practices from the perspective of hospital managers and leaders, and did not involve patient participants. The research question and outcome measures were defined based on operational indicators and institutional perceptions collected from healthcare professionals in managerial roles. Given the organisational and managerial focus of the study, patient and public involvement was not considered applicable to the research design. The absence of patient involvement does not affect the validity of the findings, which relate to institutional capacity and governance rather than patient-level outcomes.

### 2.10 Ethical Aspects

The study was conducted in compliance with national ethical guidelines and approved by the Research Ethics Committee of Hospital Sírio-Libanês (CAAE: 91286824.4.0000.5461; opinion no. 7,953,619). Participation was voluntary, with electronic acceptance of the Informed Consent Form (ICF) before beginning the questionnaire.

All responses were collected anonymously, and no patient clinical data were used. Data were stored in a secure environment and handled in accordance with the General Data Protection Law (LGPD), ensuring confidentiality and security of information.

## 3. RESULTS

### 3.1 Sample Characteristics

The questionnaire was answered by 12 of the 16 hospitals participating in Phase 2 of the Lean in Emergency Departments Project, corresponding to a response rate of 75.0% (Figure 1). The majority of respondents were nursing professionals (66.7%), followed by medicine (25.0%) and other backgrounds (8.3%). Regarding positions, managers predominated (50.0%) and directors (33.3%). Participant characteristics are detailed in Table 1.

**Table 1.**
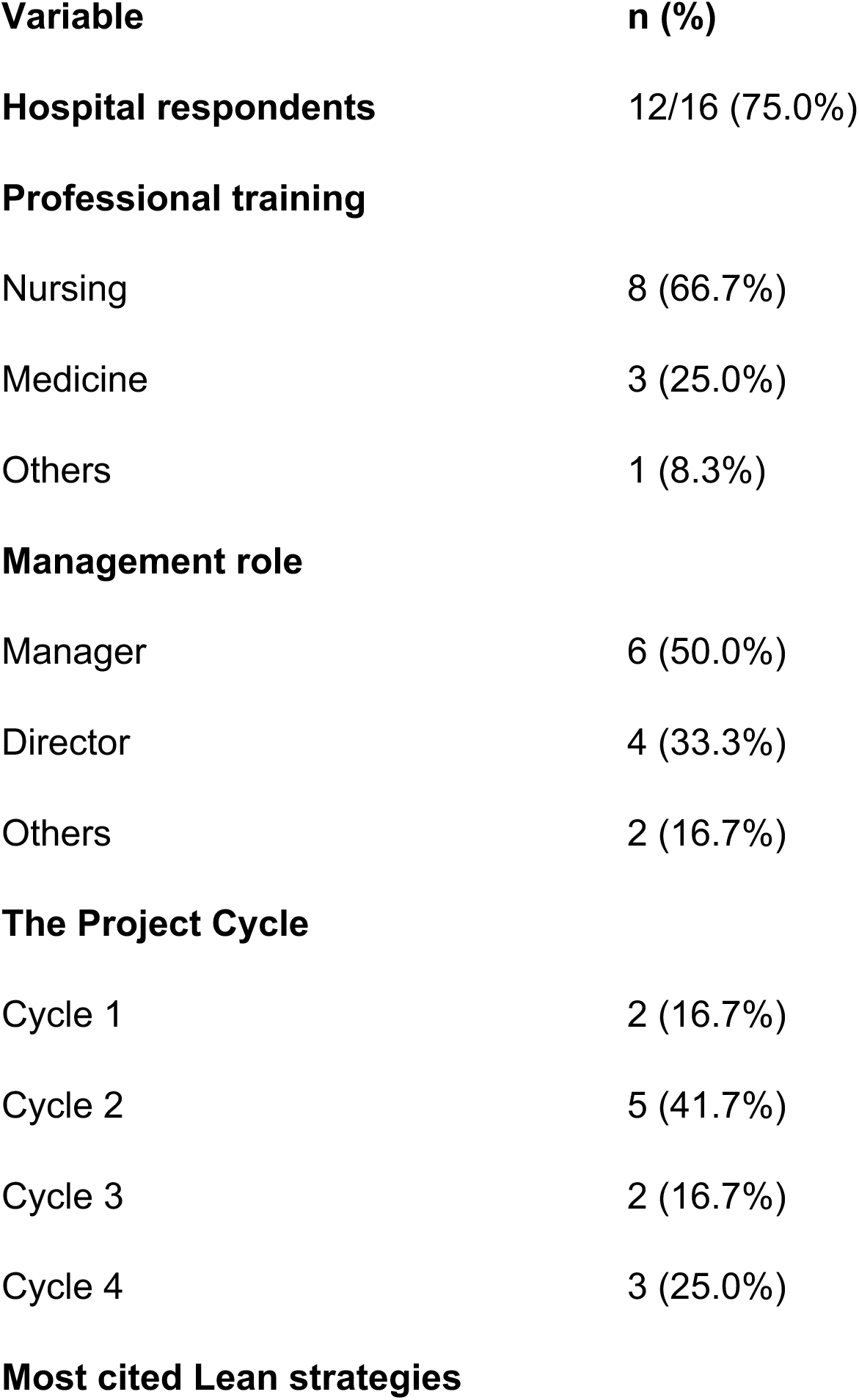
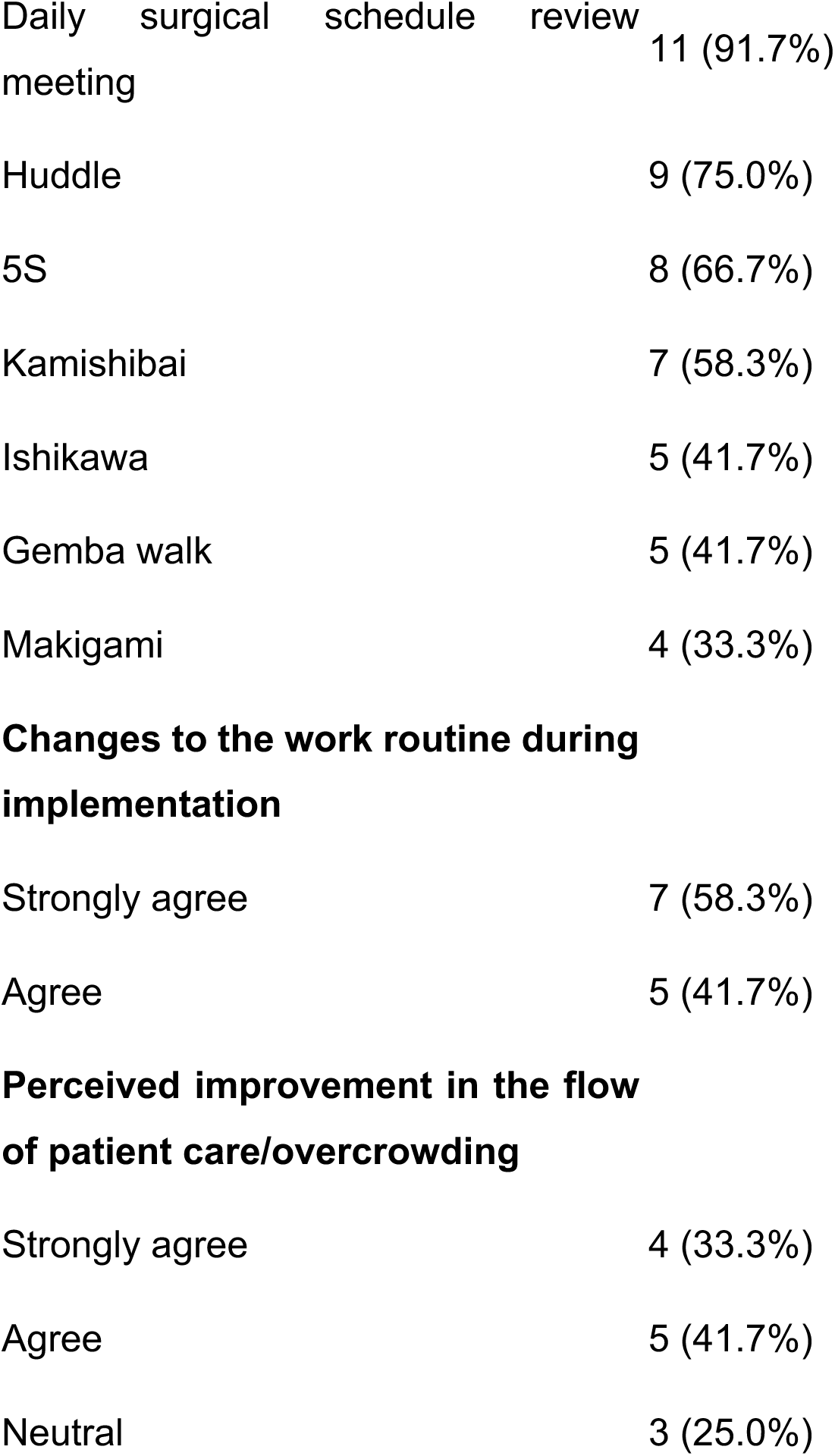
Characteristics of respondents and initial implementation findings.

| **Variable** | **n (%)** |
| --- | --- |
| **Hospital respondents** | 12/16 (75.0%) |
| **Professional training** |  |
| Nursing | 8 (66.7%) |
| Medicine | 3 (25.0%) |
| Others | 1 (8.3%) |
| **Management role** |  |
| Manager | 6 (50.0%) |
| Director | 4 (33.3%) |
| Others | 2 (16.7%) |
| **The Project Cycle** |  |
| Cycle 1 | 2 (16.7%) |
| Cycle 2 | 5 (41.7%) |
| Cycle 3 | 2 (16.7%) |
| Cycle 4 | 3 (25.0%) |
| **Most cited Lean strategies** |  |
| Daily surgical schedule review meeting | 11 (91.7%) |
| Huddle | 9 (75.0%) |
| 5S | 8 (66.7%) |
| Kamishibai | 7 (58.3%) |
| Ishikawa | 5 (41.7%) |
| Gemba walk | 5 (41.7%) |
| Makigami | 4 (33.3%) |
| **Changes to the work routine during implementation** |  |
| Strongly agree | 7 (58.3%) |
| Agree | 5 (41.7%) |
| **Perceived improvement in the flow of patient care/overcrowding** |  |
| Strongly agree | 4 (33.3%) |
| Agree | 5 (41.7%) |
| Neutral | 3 (25.0%) |

### 3.2 Implementation and Adoption of Lean Practices

The tools most frequently associated with Lean adoption were visual management boards (91.7%), huddles (75.0%), 5S (66.7%), and kamishibai (58.3%). Analytical tools, such as Ishikawa diagram and gemba walk, were less frequently cited (41.7% each) (Table 1).

All respondents reported changes in work routine during implementation, with 58.3% “strongly agree” and 41.7% “agree”. The majority (75.0%) also reported improvement in care flow and overcrowding during the intervention period (Table 1).

### 3.3 Institutional Incorporation and Barriers to Change

The alignment of professionals not directly involved in the project was considered positive by 58.3% of respondents, whilst 25.0% reported neutral perception and 16.7% negative. A similar result was observed for perception of institutional cultural transformation (58.3% positive).

The presence of resistance to change was reported by 83.3% of participants. Among the most frequently cited hindering factors were difficulties in institutional alignment (66.7%), time and effort required for implementation (58.3%), changes in previously established processes (50.0%), and uncertainties regarding cultural transformation (50.0%).

### 3.4 Sustainability of Lean Practices Over Time

Maintenance of changes after the initial implementation phase showed reduction compared to the intervention period, with 58.3% positive responses, 16.7% neutral, and 25.0% negative.

In the short term (during follow-up of up to one year), 66.7% of respondents reported maintenance of practices. In the long term, after structured follow-up ended, a similar distribution was observed between positive responses (33.3%), neutral (33.3%), and negative (33.3%) (Table 2). Although short-term sustainability was reported by most respondents (66.7%), this declined substantially after structured follow-up ended (33.3%), indicating a reduction in sustained practices over time.

**Table 2.**
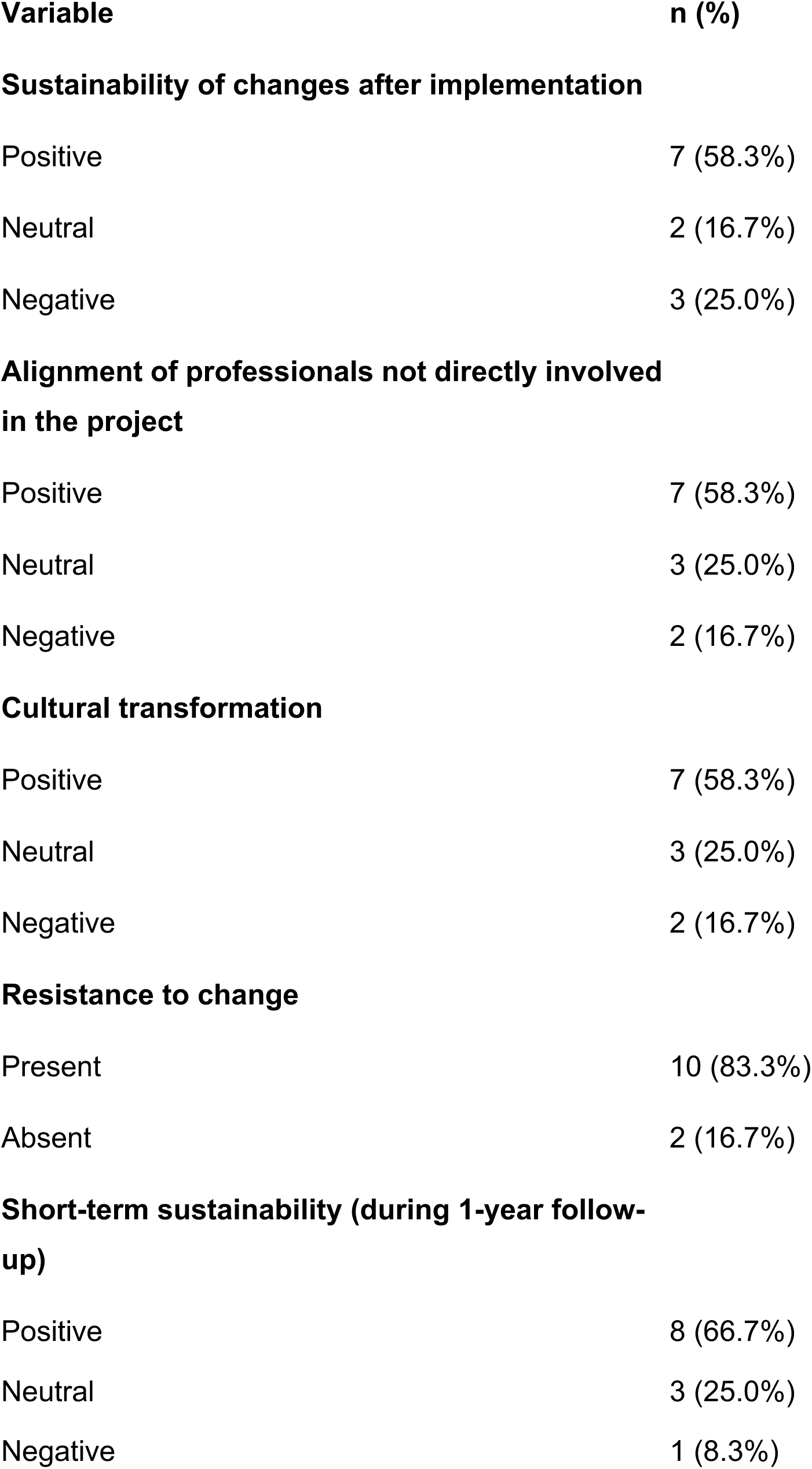
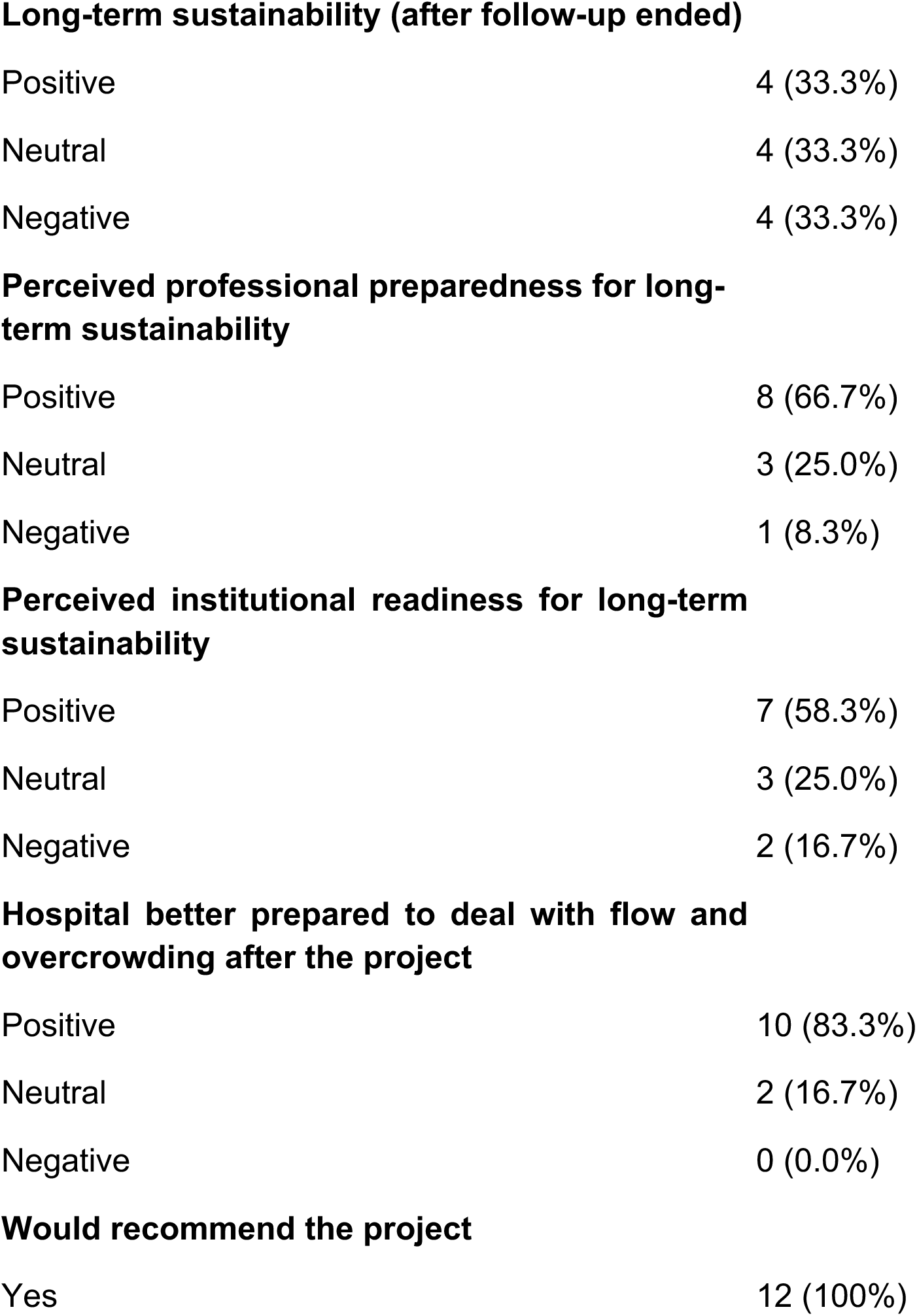
Perceptions regarding sustainability, organisational culture, and impact of Lean after implementation.

| **Variable** | **n (%)** |
| --- | --- |
| **Sustainability of changes after implementation** |  |
| Positive | 7 (58.3%) |
| Neutral | 2 (16.7%) |
| Negative | 3 (25.0%) |
| **Alignment of professionals not directly involved in the project** |  |
| Positive | 7 (58.3%) |
| Neutral | 3 (25.0%) |
| Negative | 2 (16.7%) |
| **Cultural transformation** |  |
| Positive | 7 (58.3%) |
| Neutral | 3 (25.0%) |
| Negative | 2 (16.7%) |
| **Resistance to change** |  |
| Present | 10 (83.3%) |
| Absent | 2 (16.7%) |
| **Short-term sustainability (during 1-year follow-up)** |  |
| Positive | 8 (66.7%) |
| Neutral | 3 (25.0%) |
| Negative | 1 (8.3%) |
| **Long-term sustainability (after follow-up ended)** |  |
| Positive | 4 (33.3%) |
| Neutral | 4 (33.3%) |
| Negative | 4 (33.3%) |
| **Perceived professional preparedness for long-term sustainability** |  |
| Positive | 8 (66.7%) |
| Neutral | 3 (25.0%) |
| Negative | 1 (8.3%) |
| **Perceived institutional readiness for long-term sustainability** |  |
| Positive | 7 (58.3%) |
| Neutral | 3 (25.0%) |
| Negative | 2 (16.7%) |
| **Hospital better prepared to deal with flow and overcrowding after the project** |  |
| Positive | 10 (83.3%) |
| Neutral | 2 (16.7%) |
| Negative | 0 (0.0%) |
| **Would recommend the project** |  |
| Yes | 12 (100%) |

Regarding capacity to maintain results, 66.7% of participants indicated that professionals felt capable of maintaining results, whilst 58.3% considered that the hospital was adequately structured to sustain them (Table 2).

### 3.5 Global Perception of the Impact

The majority of respondents (83.3%) considered that the hospital was better prepared to manage overcrowding and care flow after the intervention. All participants (100%) indicated that they would recommend the project to other hospitals.

Open-ended responses highlighted the importance of institutionalising Lean practices, strengthening local leadership, continuous training in the face of professional turnover, and adoption of strategies aimed at the maintenance phase.

### 3.6 Convergence Between Institutional Perception and Operational Indicators

Analysis of operational indicators demonstrated heterogeneous variation across implementation cycles (Figure 2). Delay of first elective surgery was reduced in all four cycles (Cycle 1: −50%; Cycle 2: −49%; Cycle 3: −20%; Cycle 4: −57%), representing the most consistent finding across hospitals. Operating theatre efficiency improved in three of four cycles (Cycle 2: +33%; Cycle 3: +19%; Cycle 4: +69%), but decreased in Cycle 1 (−12%). Elective surgery cancellation rate was reduced in three cycles (Cycle 2: −5%; Cycle 3: −22%; Cycle 4: −37%) but increased in Cycle 1 (+13%). Hospital length of stay showed the most variable pattern, with reductions in Cycle 1 (−10%) and Cycle 3 (−13%), a marginal change in Cycle 2 (−1%), and an increase in Cycle 4 (+9%). No consistent pattern of improvement was observed across all four indicators simultaneously in any single cycle, suggesting relevant influence of local structural conditions on intervention outcomes. The patterns observed in operational indicators accompanied the variability of institutional perceptions regarding the continuity of practices over time.

**Figure 2.**
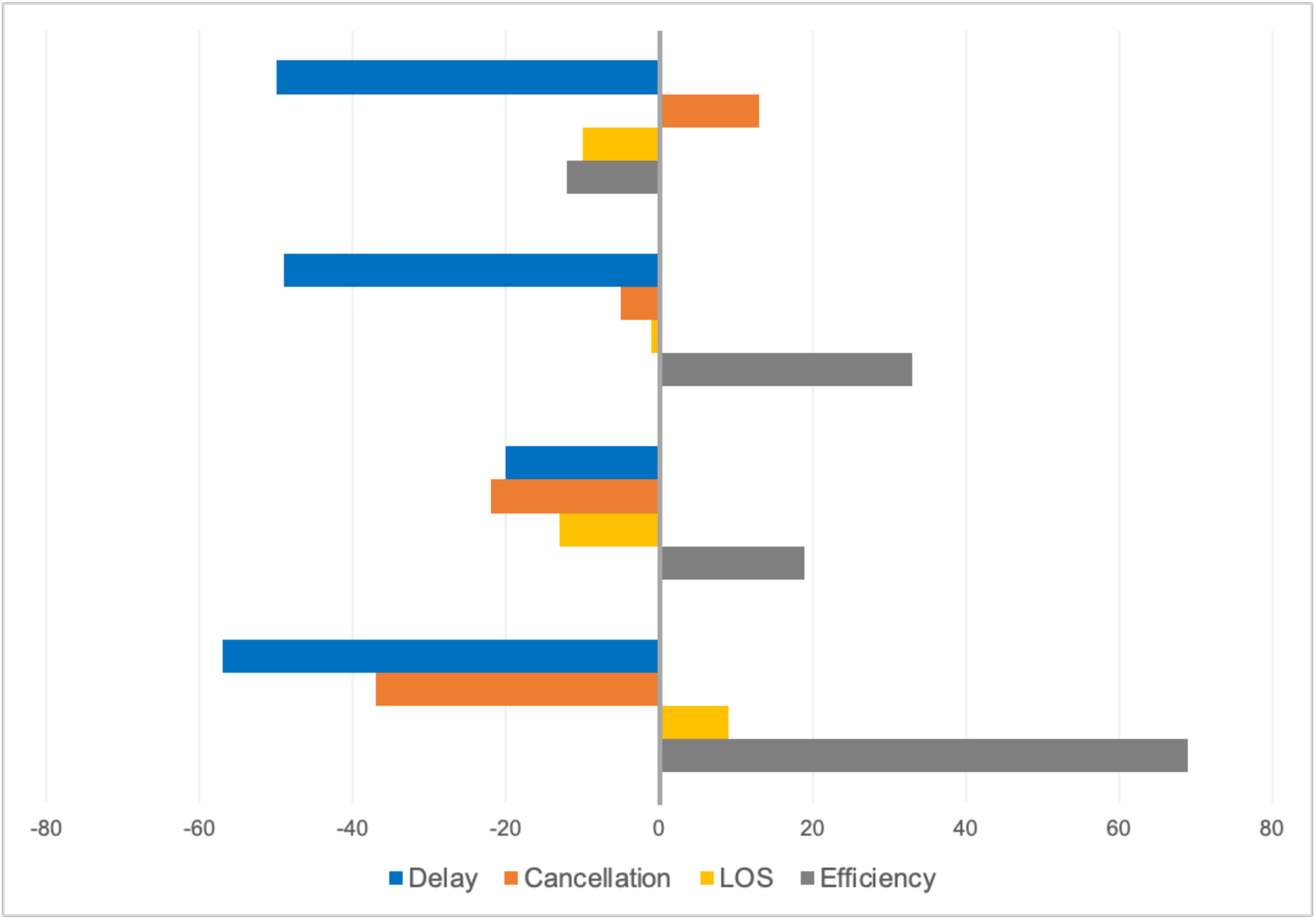
Percentage variation of operational indicators by implementation cycle. Each cycle represents a distinct group of hospitals. Bars show the percentage change in key operational indicators after the intervention, including delay of first elective surgery (Delay), elective surgery cancellation rate (Cancellation), hospital length of stay (LOS), and operating theatre efficiency (Efficiency). For Delay, Cancellation, and LOS, negative values indicate improvement (reduction), whereas for Efficiency, positive values indicate improvement. Marked heterogeneity is observed across cycles, with no consistent pattern of improvement, suggesting the influence of local organisational and structural factors on intervention outcomes.

An exploratory comparison between long-term sustainability outcomes and operational indicators by implementation cycle is presented in Table 3. Hospitals reporting positive long-term sustainability were predominantly concentrated in Cycle 2, which also showed the most balanced operational improvement profile. In contrast, all hospitals in Cycle 3 reported negative long-term sustainability despite observable improvements during the intervention. Cycle 4 presented the greatest internal heterogeneity, with hospitals reporting both neutral and negative sustainability within the cycle that registered the largest efficiency gain (+69%), suggesting that strong operational results during the intervention period do not necessarily translate into sustained practices over time.

**Table 3.** Exploratory comparison of long-term sustainability outcomes and operational indicators by implementation cycle.

| Implementation cycle | n | Positive<br>n (%) | Neutral<br>n (%) | Negative<br>n (%) | Efficiency<br>(%) | LOS<br>(%) | Cancellation<br>(%) | Delay<br>(%) |
| --- | --- | --- | --- | --- | --- | --- | --- | --- |
| Cycle 1 | 2 | 1 (50.0) | 0 (0.0) | 1 (50.0) | -12 | -10 | +13 | -50 |
| Cycle 2 | 5 | 3 (60.0) | 2 (40.0) | 0 (0.0) | +33 | -1 | -5 | -49 |
| Cycle 3 | 2 | 0 (0.0) | 0 (0.0) | 2 (100.0) | +19 | -13 | -22 | -20 |
| Cycle 4 | 3 | 0 (0.0) | 2 (66.7) | 1 (33.3) | +69 | +9 | -37 | -57 |

*Positive = score 4–5; Neutral = score 3; Negative = score 1–2 (5-point Likert scale, Q14). Operational indicators represent percentage change from baseline. Exploratory analysis — not intended for inferential purposes.*

### 3.7 Qualitative Analysis of Open-Ended Responses

Thematic analysis of open-ended responses identified eight main themes related to sustainability of Lean practices in the post-implementation period. The most frequently mentioned factor was professional and managerial turnover (7/12), described as a structural threat to continuity of practices, associated with loss of institutional memory and difficulty in incorporating the methodology by new leadership.

The need for formal governance and committed leadership (6/12) emerged as a central condition for maintaining results. Hospitals that maintained stable leadership and senior management support showed greater continuity of practices, whilst contexts with managerial discontinuity evidenced progressive deterioration of results. Active monitoring and maintenance of continuous improvement cycles (6/12) were identified as fundamental operational mechanisms for sustainability, including visual management, systematic monitoring of indicators, and periodic meetings.

Additionally, participants highlighted the transition of Lean from project to institutional practice (5/12) as a relevant challenge, associated with the need for specific training for the maintenance phase and perception of gaps in post-implementation support. Other themes included dynamic cultural resistance, with tendency for regression after structured follow-up ended, and organic diffusion of practices beyond teams directly involved. Representative examples of participant responses are presented in Supplementary Material 3.

## DISCUSSION

This study evaluated the sustainability of Lean Healthcare practices after the implementation phase of a national quality improvement programme, evidencing that maintenance of gains depends not only on initial adoption of tools, but above all on their incorporation into organisational functioning over time. The findings indicate that, although practices and routines were partially maintained, continuity of gains was associated with institutional capacity to sustain structured mechanisms of management, monitoring, and governance. These findings are consistent with previous literature that describes sustainability as a dynamic phenomenon dependent on integration of interventions into organisational systems.^9,13,14^

One of the central findings of this study was the discrepancy between perceived individual capacity and organisational readiness. Although the majority of respondents indicated that professionals felt capable of maintaining Lean practices, a smaller proportion considered that hospitals were adequately structured to sustain results in the long term. This misalignment suggests that the continuity of improvements depends less on individual competencies and more on the presence of stable organisational structures, including governance mechanisms, institutional support, formal monitoring processes, and integration of Lean practices into routine management. In this sense, sustainability emerges as an organisational property rather than an individual capability. Such misalignment appears to constitute a central explanatory mechanism for the loss of continuity of Lean interventions in real-world practice contexts, consistent with studies highlighting the role of structural conditions and institutional readiness in the implementation and maintenance of complex interventions.^9–11,15^ Qualitative analysis deepened these findings by revealing underlying mechanisms for the loss of sustainability, particularly professional and managerial turnover and the distinction between implementation and maintenance phases, the latter requiring specific training, formal monitoring mechanisms, and improvement cycles adapted to the post-project context. These findings reinforce that sustainability should not be conceptualised merely as an extension of implementation, but as a distinct phase requiring dedicated organisational capabilities.

The results reinforce the systemic nature of health interventions, in which sustainable changes require alignment between different organisational levels. The quality improvement literature demonstrates that well-executed interventions during the implementation phase may not be sustained over time in the absence of structures that ensure operational continuity, institutional prioritisation, and local adaptation.^4,9,14^ In this sense, sustainability should be understood as a dynamic phenomenon, influenced by the interaction between care practices, management processes, and institutional environment.^11,14^ The combined analysis between professional perception and operational indicators allowed a more robust understanding of findings. In some scenarios, partial maintenance of Lean practices was observed despite variations in indicators, whilst in others there was deterioration of results associated with reduced adherence to structured routines. This triangulation suggests that the mere continuation of Lean practices is insufficient to ensure stable performance over time, reinforcing the need for active management mechanisms and institutional embedding to sustain outcomes.^5,8,14^

An exploratory comparison between hospitals with positive versus negative long-term sustainability outcomes revealed potentially relevant patterns (Table 3). Hospitals reporting positive long-term sustainability were predominantly concentrated in Cycle 2, which also showed the most balanced operational improvement profile, simultaneous gains across three of four indicators without deterioration in any. In contrast, all hospitals in Cycle 3 reported negative long-term sustainability, despite observable improvements in surgical cancellation rate and length of stay during the intervention. Notably, Cycle 4 presented the greatest internal heterogeneity: hospitals with both neutral and negative sustainability outcomes were observed within the same cycle that registered the largest efficiency gain (+69%), suggesting that strong operational results during the intervention period do not necessarily translate into sustained practices over time. Qualitative responses corroborated these patterns, with hospitals reporting positive sustainability consistently citing stable leadership, structured governance mechanisms, and systematic indicator monitoring as enabling factors, while those with negative sustainability highlighted management turnover, absence of dedicated teams, and loss of institutional momentum following the end of structured support. These findings should be interpreted cautiously given the small sample size, but they generate hypotheses that warrant investigation in future longitudinal studies with larger samples.

The presence of formal governance structures, such as dedicated teams, monitoring routines, and accountability mechanisms, emerged as a central element for maintaining results, consistent with theoretical models emphasising leadership and institutionalisation of processes as determinants of sustainability.^10,11,16^ By integrating organisational perception with operational indicators in a multicentre context, this study provides a more comprehensive understanding of sustainability in Lean interventions.

These findings have important implications for the design of future quality improvement programmes. They suggest that sustainability of Lean interventions requires not only initial training and implementation support, but also the establishment of formal governance structures, continuous monitoring systems, and integration into routine management practices. Future programmes should incorporate explicit sustainability strategies from the outset, including leadership development, structured follow-up mechanisms, and institutional accountability processes. These findings may be particularly relevant for large-scale quality improvement programmes implemented in complex health systems.

Some limitations should be considered. The use of a single key respondent per hospital represents an additional limitation that warrants consideration. Although respondents were selected based on their strategic involvement in project implementation, their perceptions may not fully reflect the institutional experience as a whole, and may be subject to social desirability bias or positivity bias regarding their own organisations. This design choice may have introduced systematic overestimation of sustainability outcomes, particularly in hospitals where the respondent held a leadership role directly accountable for project results. Future studies should consider collecting data from multiple informants per institution, including frontline professionals, to enable a more comprehensive and triangulated assessment of organisational sustainability. Furthermore, heterogeneity among participating hospitals may influence results, reflecting contextual differences in implementation and maintenance of practices, which may limit generalisability to other health systems. Finally, the observational design does not allow establishing definitive causal relationships between implemented practices and observed outcomes, although it offers relevant evidence on associations in real-world practice contexts. Future research should explore longitudinal designs and intervention strategies specifically aimed at strengthening sustainability in different organisational contexts.

## CONCLUSION

In summary, the sustainability of Lean Healthcare practices depends not only on the initial adoption of tools or individual training, but fundamentally on the capacity of organisations to embed, maintain, and adapt these practices over time. These findings highlight the need to design quality improvement programmes with explicit strategies for sustainability, incorporating governance structures, continuous monitoring, and institutional accountability as core components rather than secondary considerations.

## Data Availability

The data supporting the findings of this study are not publicly available due to institutional and ethical restrictions. De-identified survey data and aggregated operational data may be available from the corresponding author upon reasonable request and subject to the necessary institutional approvals.

## Author contributions

BDDO conceived the study, conducted the analysis, and drafted the manuscript. MSB contributed to the study design and critical revision of the manuscript. WGRP contributed to data collection and interpretation of the results. PAR and CTP contributed to the study conception and critical revision of the manuscript for important intellectual content. All authors reviewed, approved the final version of the manuscript, and agree to be accountable for all aspects of the work.

## Data availability statement

The anonymised dataset, study questionnaire, and qualitative responses supporting the findings of this study are openly available at the Open Science Framework via Zenodo: https://doi.org/10.5281/zenodo.19635843

