## Supplementary Material for "Sustainability of Lean interventions in public hospitals after a national quality improvement programme: a multicentre mixed-methods study"

**Supplementary Material 1.** Methodological framework for Lean Healthcare interventions throughout the implementation and monitoring phases

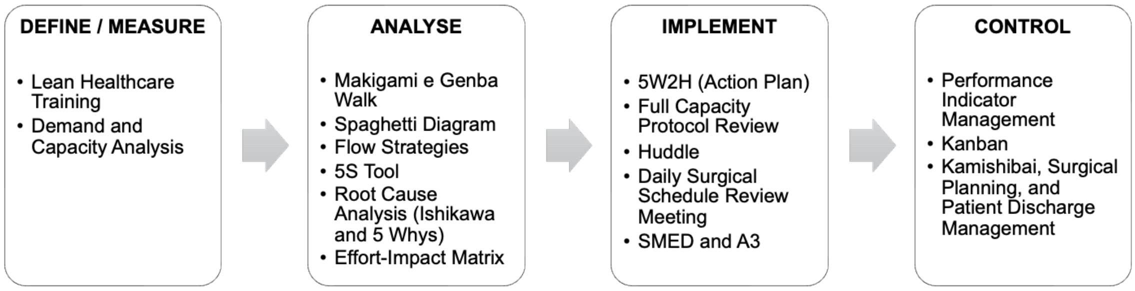


Source: Adapted from the Lean in Emergencies Project model (Brazilian Ministry of Health / PROADI-SUS).

The figure presents the methodological structure of Lean Healthcare-based interventions, organised into four macro stages: define/measure, analyse, implement, and control. In each phase, the main tools used in the hospital context are highlighted, including demand and capacity diagnosis, process analysis (Makigami, Gemba Walk, and spaghetti diagram), development and implementation of action plans (5W2H, huddles, and process review), as well as monitoring and control mechanisms such as indicator management, Kanban, and visual audits (Kamishibai). The model illustrates the integration between flow analysis, decision-making, and continuous operational management.

**Supplementary Material 2.** Questionnaire

1. Professional background:

    a. Nursing
    b. Medicine
    c. Administration
    d. Other

2. Management position:

    a. Director
    b. Superintendent
    c. Manager
    d. Area Coordinator/Supervisor
    e. Unit head
    f. Division head
    g. Sector head
    h. Not applicable

3. Participant cycle:

    a. Cycle 1
    b. Cycle 2
    c. Cycle 3
    d. Cycle 4

4. During the implementation of Lean in Emergencies, which strategies most enhanced the adoption of Lean in your hospital? Select up to 5 options.

    a. Makigami
    b. Spaghetti diagram
    c. 5S
    d. Ishikawa diagram
    e. Gemba walk

f. Huddle
g. Process mapping
h. SMED
i. Kamishibai

5. DURING the implementation period of Lean in Emergencies, there was a clear change in the daily work routine of the professionals in our hospital because of the project.

   a. Strongly agree
   b. Agree
   c. Neither agree nor disagree
   d. Disagree
   e. Strongly disagree

6. AFTER the completion of Lean in Emergencies, the changes in the daily work routine of the professionals in our hospital were MAINTAINED.

   a. Strongly agree
   b. Agree
   c. Neither agree nor disagree
   d. Disagree
   e. Strongly disagree

7. Even professionals in our hospital who were not directly involved with Lean in Emergencies aligned themselves with Lean.

   a. Strongly agree
   b. Agree
   c. Neither agree nor disagree
   d. Disagree
   e. Strongly disagree

8. There was a clear cultural transformation in our hospital as a result of Lean in Emergencies.

a. Strongly agree
b. Agree
c. Neither agree nor disagree
d. Disagree
e. Strongly disagree

9. There was resistance from certain teams/individuals to the changes brought about by Lean.

    a. Strongly agree
    b. Agree
    c. Neither agree nor disagree
    d. Disagree
    e. Strongly disagree

10. If applicable, indicate the main points of resistance or challenges your hospital encountered during the project implementation. Select up to 3 options.

    a. Establishing effective communication
    b. Aligning the entire hospital team
    c. Uncertainty about how Lean would affect personal work routines
    d. Uncertainty about how Lean would affect the work dynamics between individuals or teams
    e. Uncertainty about how Lean could change processes that had long been practised and accepted
    f. Uncertainty about the feasibility of the cultural transformation in the hospital required for Lean success
    g. Uncertainty about the time and effort required to implement Lean
    h. Uncertainty about how the project's long term results would be maintained

11. Note: before proceeding, would you like to make any observation/comment about your answers regarding changes in work

routines, cultural transformation, or team alignment/resistance? Please tell us below.

12. During the 6-month intervention period of Lean in Emergencies, our hospital observed improvements in emergency overcrowding and workflow management.

    a. Strongly agree
    b. Agree
    c. Neither agree nor disagree
    d. Disagree
    e. Strongly disagree

13. SHORT TERM: during the 1 year follow up period, our hospital was able to maintain the sustainability of the Lean methodology and culture.

    a. Strongly agree
    b. Agree
    c. Neither agree nor disagree
    d. Disagree
    e. Strongly disagree

14. LONG TERM: after the end of the 1 year follow up period, our hospital was still able to maintain the sustainability of the Lean methodology and culture.

    a. Strongly agree
    b. Agree
    c. Neither agree nor disagree
    d. Disagree
    e. Strongly disagree

15. Thanks to the training provided by the Lean in Emergencies team, the professionals in our hospital who participated in the project feel prepared to maintain the project’s results in the long term.

a. Strongly agree
b. Agree
c. Neither agree nor disagree
d. Disagree
e. Strongly disagree

16. Our hospital is equipped to maintain the project’s results in the long term.

    a. Strongly agree
    b. Agree
    c. Neither agree nor disagree
    d. Disagree
    e. Strongly disagree

17. What are the main CHALLENGES your hospital is facing to keep the sustainability of the Lean methodology and culture alive and active in the long term?

18. What MAIN STRATEGIES is your hospital adopting to keep the sustainability of the Lean methodology and culture alive and active in the long term?

19. Note: before proceeding, would you like to make any observation/comment about your answers regarding sustainability, team training, or maintenance of short and long term results? Please tell us below.

20. Which do you consider to be the strengths of the Lean in Emergencies project? Select all relevant options.

    a. Lean training
    b. Team training
    c. Identification of improvement opportunities
    d. Result monitoring
    e. Capacity and demand analysis
    f. Method for building the action plan

g. Emergency services
h. Change management (cultural transformation)

21. What do you think we could improve? Please be specific.

22. Our hospital is better prepared to deal with overcrowding and workflow management after Lean in Emergencies compared with before the intervention.

    a. Strongly agree
    b. Agree
    c. Neither agree nor disagree
    d. Disagree
    e. Strongly disagree

23. Would you recommend other hospitals to enrol in Lean in Emergencies?
    a. Yes
    b. No

24. Was there anything that surprised you about the implementation of Lean in Emergencies?

25. Is there anything else you would like to inform our team?

**Supplementary Material 3.** Illustrative examples of qualitative responses

This material presents representative excerpts from the open responses provided by participants, organised by themes identified in the thematic analysis. The quotations were anonymised and selected to illustrate the main factors related to the sustainability of Lean practices in the post implementation period. Quotations were selected and, where necessary, paraphrased to prevent identification of individual respondents or institutions, in accordance with the anonymity procedures described in the Methods section.

Theme 1. Staff turnover and workforce instability

- “A significant proportion of the team was replaced during the post-implementation period.”
- “Training teams because of turnover.”
- “The arrival of new professionals changes the work dynamics.”

Theme 2. Governance and institutional leadership

- “A strong leader is needed, with support from senior management.”
- “Strengthening the leadership role and the teams.”
- “Management support for the project tools is fundamental.”

Theme 3. Monitoring and management routines

- “Maintaining huddles, Kanban, and discharge room routines.”
- “Systematic monitoring of indicators.”
- “Visual management with periodic meetings.”

Theme 4. Resistance to change and organisational culture

- “There was a lot of resistance, especially from medical teams.”
- “People resist stepping out of their comfort zone.”
- “People always return to their comfort status.”

Theme 5. Challenges in post implementation sustainability

- “Maintaining changes is challenging.”
- “Good practices are lost if there is no institutionalisation.”
- “After the projects end, maintaining standards over time is difficult.”

Theme 6. Institutionalisation and continuous improvement

- “Consolidating Lean thinking as a daily practice, not just as a one-off project.”
- “Lean culture requires discipline, consistency, and engagement.”
